# Expanding the phenotypic spectrum of TRAF7 syndrome: report of eleven new cases and literature review

**DOI:** 10.1101/2023.12.13.23299272

**Authors:** Carmen Palma-Milla, Aina Prat-Planas, Emma Soengas-Gonda, Mónica Centeno-Pla, Jaime Sánchez-Pozo, Irene Lazaro-Rodriguez, Juan F. Quesada-Espinosa, Ana Arteche-Lopez, Jonathan Olival, Marta Pacio-Miguez, María Palomares-Bralo, Fernando Santos-Simarro, Ramón Cancho-Candela, María Vázquez-López, Veronica Seidel, Antonio F Martinez-Monseny, Didac Casas-Alba, Daniel Grinberg, Susanna Balcells, Mercedes Serrano, Raquel Rabionet, Miguel A. Martin, Roser Urreizti

## Abstract

TRAF7 syndrome, a multisystemic neurodevelopmental disorder caused by germline missense variants in the *TRAF7* gene, exhibits heterogeneous clinical presentations. We present a detailed description of eleven new TRAF7 syndrome cases, featuring eight distinct variants, including a novel one. Phenotypic analysis and a comprehensive review of the 58 previously reported cases outline consistent clinical presentations, emphasizing dysmorphic features, developmental delay, endocrine manifestations, and cardiac defects. In this enlarged collection, novelties include a wider range of cognitive dysfunction, with some individuals exhibiting normal development despite early psychomotor delay. Communication challenges, particularly in expressive language, are prevalent, necessitating alternative communication methods. Autistic traits, notably rigidity, are observed in the cohort. Also, worth highlighting are hearing loss, sleep disturbances, and endocrine anomalies, including growth deficiency. Cardiac defects, frequently severe, pose early-life complications. Facial features, including arched eyebrows, contribute to the distinct gestalt. A novel missense variant, p.(Arg653Leu), further underscores the complex relationship between germline *TRAF7* variants and somatic changes linked to meningiomas. Our comprehensive analysis expands the phenotypic spectrum, emphasizing the need for oncological evaluations and proposing an evidence-based schedule for clinical management. This study contributes to a better understanding of TRAF7 syndrome, offering insights for improved diagnosis, intervention, and patient care.

**Key Message:** TRAF7 syndrome, a complex neurodevelopmental disorder, exhibits variable cognitive outcomes, complex behavioral presentation and clinical complications emphasizing the need for tailored interventions and follow-up.

## INTRODUCTION

TRAF7 syndrome (also known as CAFDADD; Cardiac, facial, and digital anomalies with developmental delay; MIM #618164; ORPHA: 592570) is a multisystemic neurodevelopmental disorder caused by heterozygous germline pathogenic variants in the *TRAF7* gene (Tumor necrosis factor receptor-associated factor 7; MIM *606692). Since its initial description in 2018 [1] and further delineation [2], a total of 58 subjects with TRAF7 syndrome have been published, carrying 26 different variants [3–8]. While the clinical presentation of the syndrome appears to be highly variable, some common features have been identified, including a range of neurological and developmental abnormalities, such as intellectual disability, speech and language delays, and motor impairments, as well as epilepsy in some individuals. Dysmorphic facial features (blepharophimosis, hypertelorism and micrognathia among others) and cardiovascular abnormalities are also seen in most individuals with TRAF7 syndrome [2]. Additionally, skeletal anomalies like digital anomalies, scoliosis or other spine deformities have been reported [1,2].

In humans, the tumor necrosis factor receptor (TNF-R)-associated factor (TRAF) family consists of seven genes, *TRAF7* being the last member to be identified. All of them share a domain organization consisting of a modular structure, which allows them to convey signals from different receptors, but each plays a unique and well-defined role in cellular biology. Specifically, TRAF7 is a multifunctional intracellular protein that acts as a key mediator of the nuclear factor-κB (NF-κB), the mitogen-activated protein kinases (MAPKs) and extracellular signal-regulated kinase (ERK) signaling pathways, playing a crucial role in the control of cell survival, apoptosis, proliferation, and differentiation [9–14].

The *TRAF7* gene encodes a 670 amino acid protein (the E3 ubiquitin-protein ligase TRAF7) containing an N-terminal RING finger domain, an adjacent TRAF-type zinc finger domain, a centrally situated coiled-coil motif, and seven WD40 repeats in the C-terminal domain. The WD40 repeats are unique to TRAF7 within the TRAF family, with all other members containing a C-terminal TRAF domain instead [10]. TRAF7 syndromic variants are germline pathogenic missense variants that cluster mostly at the WD40 repeats of the TRAF7 protein, some of them being recurrent (p.Arg655Gln, p.Arg524Trp and p.Phe617Leu being the most frequently identified).

Somatic *TRAF7* variants have also been described in a wide range of tumors: meningiomas, mesotheliomas, intraneural perineuriomas, and adenomatoid tumors of the genital tract, usually in concurrence with *KLF4, AKT1* or *PIK3CA* variants [11,15–19]. In these tumors missense variants affecting the WD40 domain are predominant, but loss-of-function variants are also present. So far, a large number of somatic cancer-related variants and an increasing number of germline syndromic-associated variants have been described, mostly overlapping in type and distribution, even affecting the same residue. However, the exact same variants have not been identified in both contexts, with the exception of a recently reported individual with mild syndromic features presenting with multiple meningiomas and a variant changing Arg641 residue to cysteine in a mosaic status [6].

Here, we report eleven subjects with a clinical and molecular diagnosis of TRAF7 syndrome, harboring *de novo* germline variants in *TRAF7*, one of which is novel. In addition, we conducted an extensive review of the literature, which allowed us to expand the clinical and genetic delineation of TRAF7 syndrome. Based on this analysis, we have developed recommendations which may contribute to a better clinical management of this ultra-rare syndrome.

## METHODS

### Subjects

Eleven individuals from different Spanish hospitals with a confirmed diagnosis of TRAF7 syndrome have been included in this study. They were recruited through a nationwide collaboration focused on TRAF7 syndrome, fostered by the CIBERER consortium [20]. The study has been conducted in accordance with the Declaration of Helsinki and has been reviewed and approved by the ethics committee at each institution. All adult participants or guardians of minors or incapacitated subjects provided written informed consent for participation.

### Literature review

We performed a Cochrane Library search and a PubMed database search from the date of the first clinical description [1] of pathology associated with variants in *TRAF7* until September 15^th^ 2023 using the terms: “TRAF7” and “CAFDADD”. In addition, the HGMD Professional 2023.3 database has been consulted covering the same period. Data extraction included first author, publication year, molecular data (identified variant and residue changes, *de novo* or inherited condition), pregnancy and perinatal information, multiorganic clinical data, complementary exams information, neuroimaging, and age and cause of death.

### Clinical Assessment

Anthropometric measures and a comprehensive dysmorphological physical examination were performed. These included the individual’s facial features, length, symmetry and presence of any abnormalities in the neck, upper and lower limbs, back, chest and cardiopulmonary function. Palpation of the abdomen for abnormalities, comprehensive genital assessment including pubertal stage and a comprehensive skin examination for cutaneous manifestations or abnormal pigmentation patterns were also performed. Cardiac anomalies, visual or auditory impairment as well as available brain neuroimages were assessed. Neurodevelopmental milestones were evaluated with Denver Developmental Screening Test [21]. Specific Autism Diagnostic Observational Schedule (ADOS-2) was addressed in patients with autistic behavior. Sleep disorders were assessed by the SDSC scale [22]. Findings were encoded using Human Phenotype Ontology codes (HPO) [23].

### Genetics

Informed consent was obtained from the patient’s parents according to International Ethical Guidelines and Declaration of Helsinki.

Whole Exome Sequencing (WES) was performed in multiple Spanish reference centers (Hospital 12 de Octubre, Hospital la Paz, Hospital Gregorio Marañón, Madrid; Hospital Sant Joan de Déu, Barcelona) according to established protocols and bioinformatic analyses following current recommendations. Variant interpretation was made according to ACMG guidelines [24]. All variants were validated and segregated by Sanger sequencing and are referenced to the canonical transcript ENST00000326181.11 (NM_032271.3). Patient 10 was previously published as Patient 33 in Castilla-Vallmanya *et al.* [2].

## RESULTS

### Clinical findings

Eleven individuals (8 male and 3 female) were recruited, ranging in age from one month to 26 years. Their clinical description is summarized in Table 1. Detailed description of each subject is available on demand. Most individuals presented unspecific prenatal complications such as single umbilical artery (3/11), polyhydramnios (3/11) or intrauterine growth retardation (2/11).

**Table 1.**
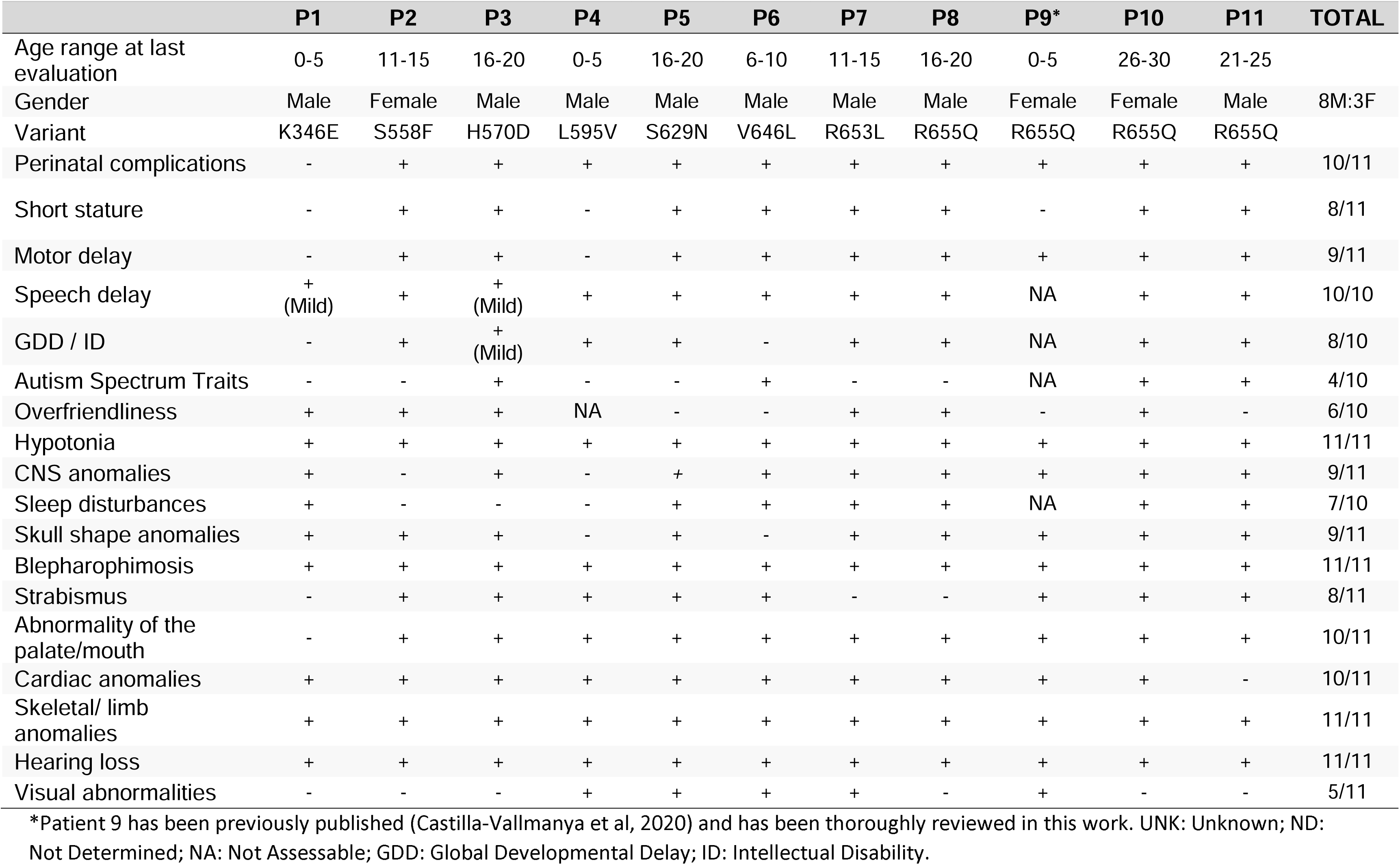
Clinical overview of the 11 patients presented here. Detailed information and HPO codes are presented in Suppl. Table 1.

#### Neurologic features

At birth, all patients showed hypotonia (11/11) with poor sucking (7/11), some of them requiring a feeding tube. Global developmental delay was common (8/10), ranging from mild to moderate. Most patients in our cohort exhibited delayed language acquisition and development (9/10), with expressive language being more severely affected than receptive language. Despite some improvement over time, all presented with some communicative difficulties. Currently, subjects P4 and P10 are communicating by sign language. Regarding neurodevelopmental comorbidities, autistic traits, lack of cognitive flexibility or emotional disorders were present in half of the cases (5/10), with two patients fulfilling Autism Spectrum Disorder (ASD) criteria evaluated by ADOS-2. Concerning behavioral phenotype, they were commonly described as having a happy demeanor or overfriendliness (7/10).

On brain magnetic resonance imaging (MRI), most of the patients (9/11) presented with structural findings such as ventriculomegaly, periventricular cysts or hydrocephalus, with no other characteristic features Epilepsy was present in only one patient (Patient 10), it appeared in the second month of life and it was well controlled with monotherapy.

As in other neurodevelopmental conditions, some kind of sleep disorder was reported for most of the patients (7/10) as assessed by SDSC scale. The most frequent were sleep-wake transition disorders (5/10), requiring medication in some patients, followed by difficulties with initiating or maintaining sleep (3/10) and disorders of excessive somnolence (3/10). Two of the patients showed abnormalities in three of the evaluated sleep areas.

#### Dysmorphic features

Dysmorphic features were common in all cases, with facial features reminiscent of TRAF7 syndrome patients (A figure with the facial features of patients identified in this study is available on demand). Skull shape anomalies were frequent but heterogeneous (9/11), trigonocephaly being the most common. Ocular examination revealed notable abnormalities with ptosis (11/11), blepharophimosis (11/11), hypertelorism (11/11) and epicanthal folds (9/11) being the most characteristic. Ear abnormalities were common but inconsistent, with low set position being the most common finding (10/11). Abnormalities of the nose were present in all patients, consisting of a round nasal tip with a prominent columella. Abnormalities of the palate/mouth were frequently seen, including micrognathia and/or retrognathia (9/11) and narrow, high or cleft palate (9/10). Dental anomalies were present in all the five patients assessed, these anomalies included malpositioned teeth and oligodontia. Broad and/or short neck were also common (10/11). Other dysmorphic features included hypoplastic, low-set or widely spaced nipples (6/10) and hypospadias in half of the male cases (4/8).

#### Cardiac features

All patients except Patient 11 presented with congenital heart defects (10/11), the most frequent cardiac malformation being patent ductus arteriosus (6/11). Other cardiac defects included aortic coarctation (4/11) and valvular or septal defects (ventricular septal defect; 4/11), bicuspid aortic valve (3/11), atrioventricular canal and atrial septal defect (1/11). Additionally, three patients presented a persistent left superior vena cava and another two, peripheral pulmonary artery stenosis. Aside from congenital heart defects, four patients had cardiac insufficiency (4/11) and four had syncopes of unknown origin (4/11).

#### Skeletal features

Digital anomalies were detected in all patients with substantial variability in the types of anomalies observed (A figure with the anomalies of the extremities of patients identified in this study is available on demand). Clinodactyly was present in five cases (5/11) and brachydactyly in four cases. Other digital anomalies included overlapping toes (4/11), syndactyly (3/11), and metatarsus adductus (3/11). Furthermore, a single patient displayed arachnodactyly.

Pectus carinatum was present in most patients (7/11). Scoliosis (6/11) and kyphosis (4/11) were also present in a range of patients. Additionally, five patients had pes planus and valgus and one had pes cavus. Two patients had 11 pairs of ribs and in two patients a marfanoid habitus was described. Joint hypermobility (4/10) or joint contractures (3/10) were also present in some cases.

#### Endocrinological features

Most patients had normal anthropometric parameters at birth, but all of them tended to lower body mass indexes during their subsequent growth (Supplementary Figure 1). Short stature and low weight were common, and most patients presented values of height below -2SD (8/11) (Figure 1), despite presenting normal IGF-1 and IGFBP-3 hormonal levels when they were analyzed. This trait is more notable if mid-parental height is considered (A detailed clinical description of the patients is available on demand). Bone age has been assessed in 4 patients and 2 showed delayed bone age (2/4). Bone mineral density has been assessed in only one case (Patient 10, at 23 years of age), and lumbar spine osteopenia (T-score of -1.3) and femoral neck osteoporosis (T-score of -2.5) were noted.

**Figure 1.**
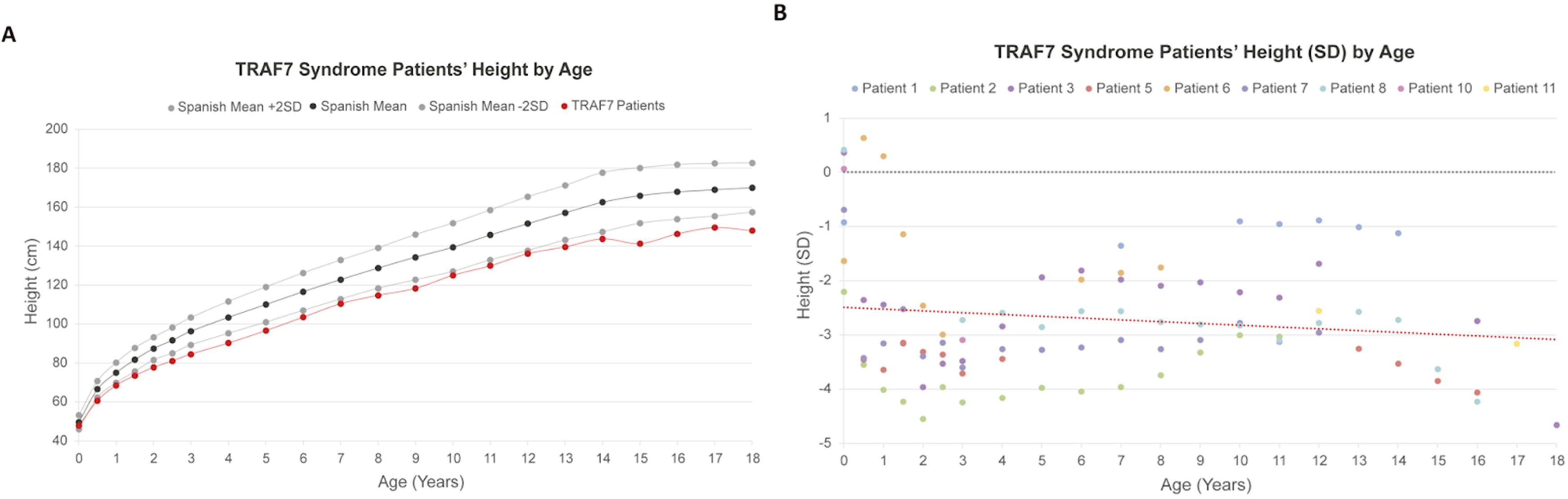
TRAF7 Syndrome Patients’ Height by Age. A) Plot of the mean height of the 9 Spanish patients reported here (red) in comparison with the Spanish population mean (average of boys and girls; black) and +/-2 SD (grey). B) Plot of the height SD by age of the 9 Spanish patients corrected by sex. The red line indicates the mean SD for all 9 TRAF7 Syndrome patients represented here. Different patients are represented in different colours. Patient 4 is excluded here due to his origin (Bulgaria) and Patient 9 is excluded due to the early age at death (9 mo).

#### Other symptoms

All patients presented hearing loss attributed to diverse etiologies: conductive (3/11), sensorineural (3/11) and mixed (3/11). Visual abnormalities were also common (9/11), including strabismus in most patients (8/11), myopia (3/11) and optic nerve atrophy (2/11).

Six patients had digestive problems, including gastroesophageal reflux and dysphagia (three patients needed feeding through a nasogastric tube or a gastrostomy). Two patients presented hepatomegaly. Five patients had renal problems and two presented hernias. Two patients had recurrent respiratory infections and one recurrent urinary tract infections.

### Proposed guidelines for the clinical management of TRAF7 syndrome

Following a thorough clinical revision of the individuals presented in this study and a systematic analysis of the 57 previously published subjects with TRAF7 syndrome (Table 2), we have developed a set of guidelines for the clinical management of this condition (Figure 2<u>*</u>), highlighting the relevance of a multidisciplinary and personalized assessment and follow up. Considering the recent publication of the first patient with a mosaic variant in *TRAF7* who developed a meningioma [6], the presence of meningiomas in one adult case (43 years old) [1] and one endometrioid adenocarcinoma in a 36 year old woman [2], oncological controls should also be considered for TRAF7 syndrome patients, especially after puberty.

**Figure 2.**
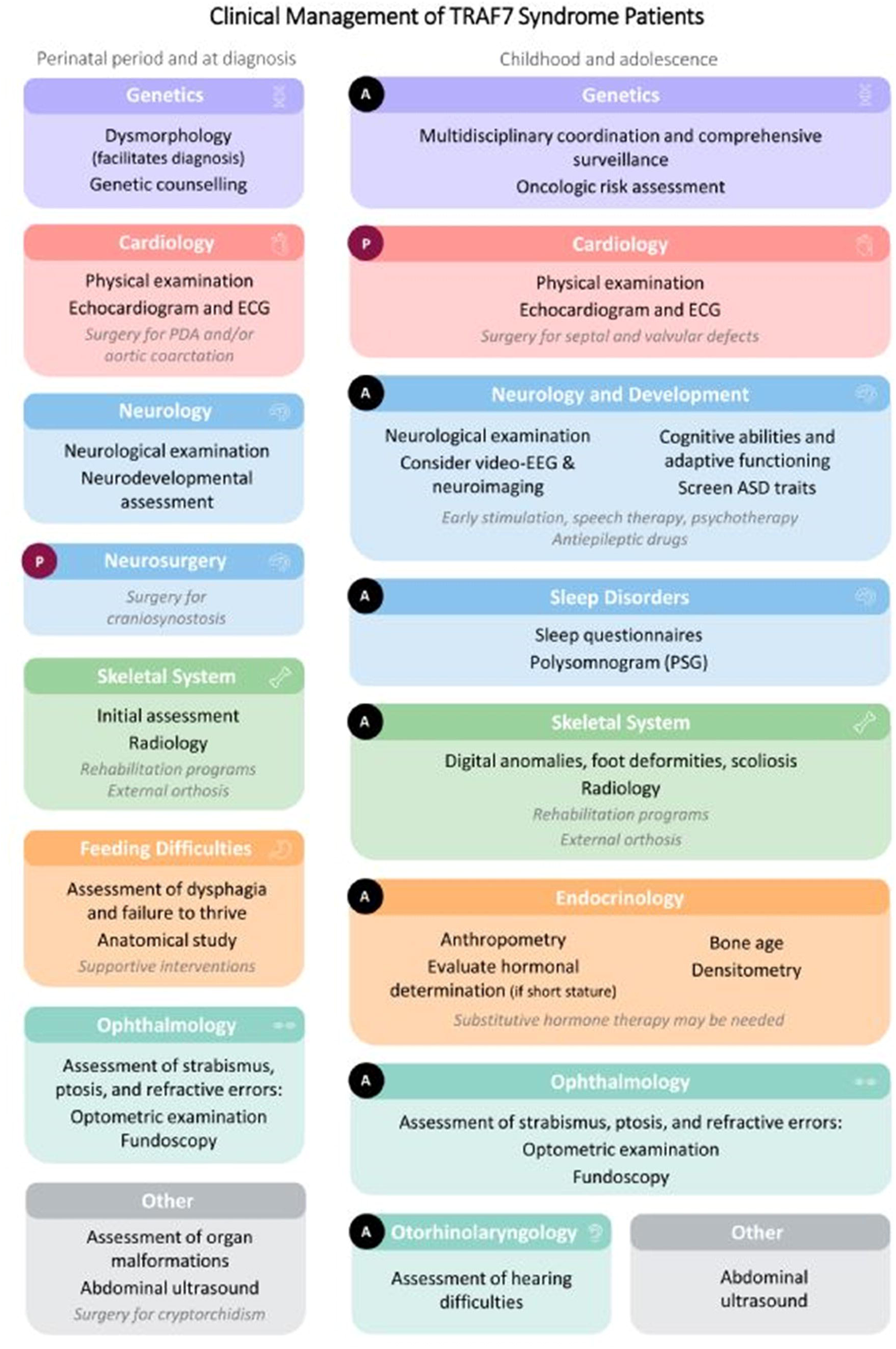
Schematic guidelines for TRAF7 syndrome clinical management. Perinatal period: Boxes include the medical area of disease. Medical problems and management requirements are detailed below. Therapies are indicated in italics. Childhood and adolescence: Boxes include medical area of disease. Medical problems, management recommendations, and complementary examinations are detailed below. Therapies are indicated in italics. An A in a black dot states that the recommended periodicity for clinical evaluation is annually, for every specialty and for all the individuals. In contrast, a Pin a burgundy dot means that those complementary examinations and management tools need to be personalized, and applied only if patient’s personal characteristics require them, always following the medical team criteria. ASD, autism spectrum disorder; EEG, electroencephalogram; ECG, electrocardiogram; PDA, patent ductus arteriosus; PSG, polysomnography. A Spanish version of this figure is available on demand.

**Table 2.**
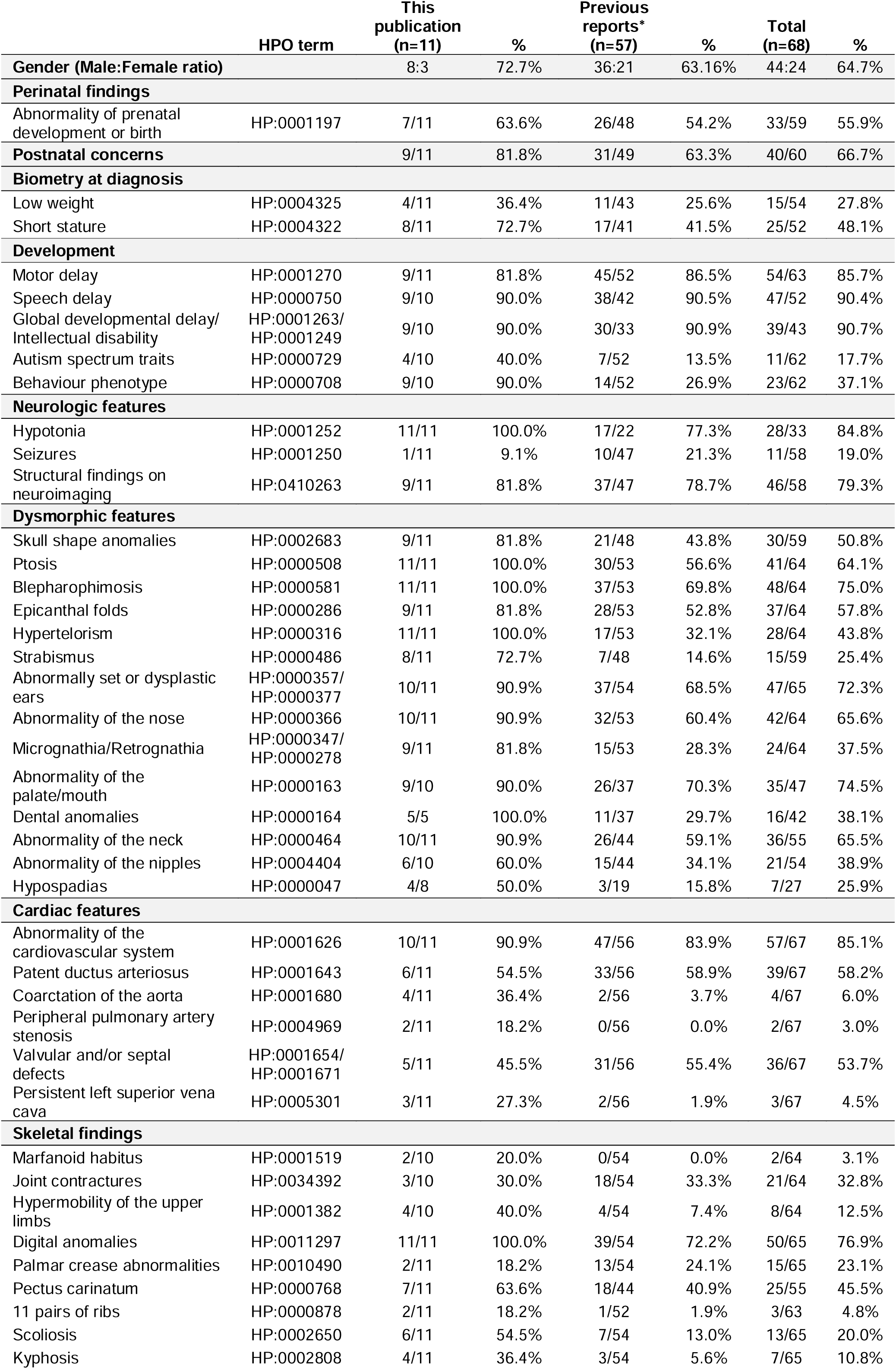
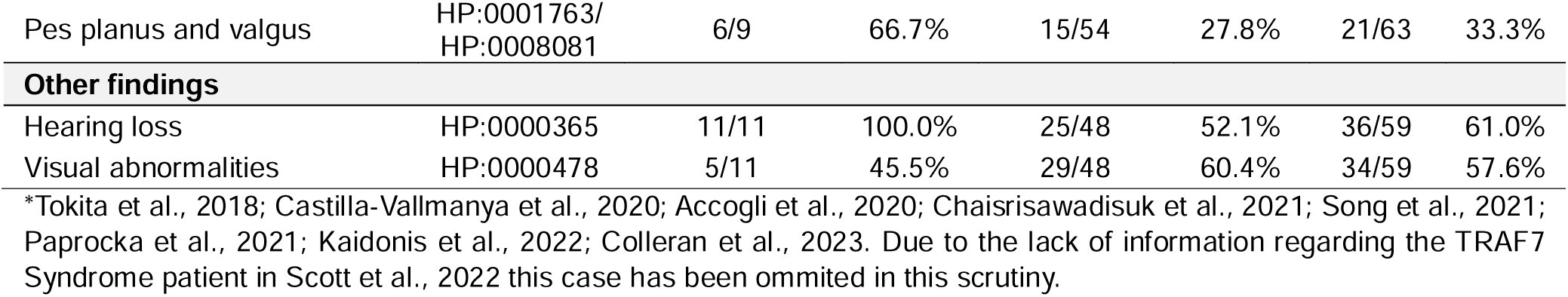
Clinical summary of all TRAF7 syndrome cases.

*A Spanish version of this figure is available on demand.

### Genetic findings

All patients are heterozygous carriers of pathogenic missense variants in *TRAF7*. All variants were confirmed to have occurred *de novo*, except for P1, conceived by ovodonation, for whom the paternal sample was negative. Eight different variants have been detected (Table 3), seven of them previously associated with TRAF7 syndrome [1,2,5,7] including the recurrent variant p.Arg655Gln, which was detected in four unrelated patients (Patients 8 to 11). Patient 7 harbors a novel variant, c.1958G>T, p.(Arg653Leu), in a residue where other amino acid substitutions have been previously reported in meningiomas [25]. As in previous studies, all variants are localized in the WD40 domains at the C-terminus of TRAF7, except for variant c.1036A>G [p.(Lys346Glu)], which affects a residue in the coiled coil domain. None of these variants were detected in populational databases (gnomAD v4; accessed November 2023), all of them affect evolutionary conserved residues and are predicted to have deleterious effects by several *in silico* tools and, with the exception of variant c.1958G>T [p.(Arg653Leu)], all have been previously identified in TRAF7 syndrome patients. This novel variant is classified as pathogenic according to ACMG criteria [24] (PS2, PM1, PM2, PP2, PP3).

**Table 3.**
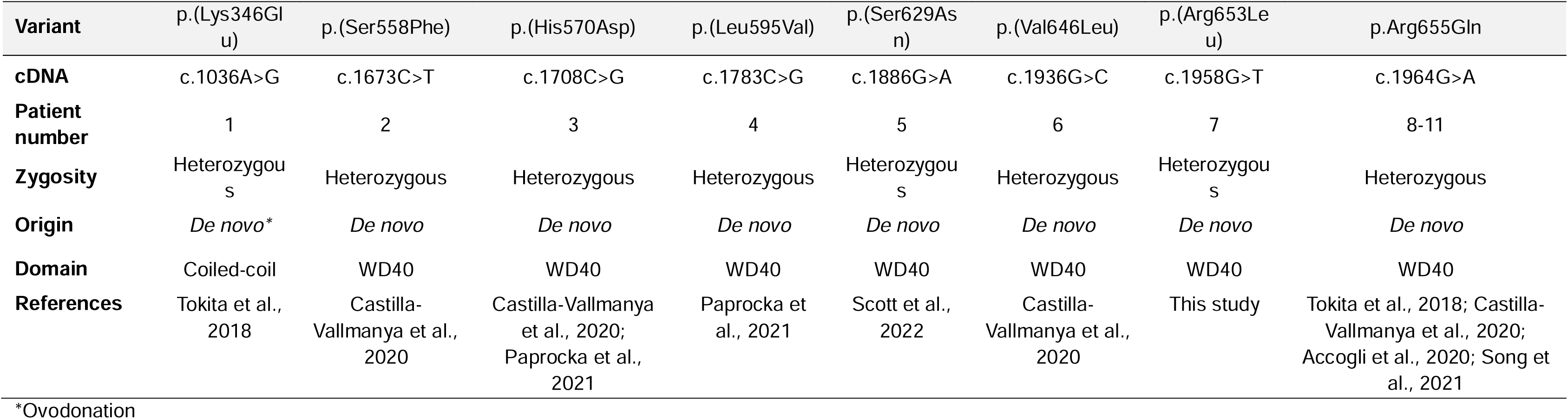
Molecular data of individuals with TRAF7 variants identified in this study.

| Variant | p.(Lys346Glu) | p.(Ser558Phe) | p.(His570Asp) | p.(Leu595Val) | p.(Ser629Asn) | p.(Val646Leu) | p.(Arg653Leu) | p.Arg655Gln |
| --- | --- | --- | --- | --- | --- | --- | --- | --- |
| **cDNA** | c.1036A>G | c.1673C>T | c.1708C>G | c.1783C>G | c.1886G>A | c.1936G>C | c.1958G>T | c.1964G>A |
| **Patient number** | 1 | 2 | 3 | 4 | 5 | 6 | 7 | 8-11 |
| **Zygosity** | Heterozygous | Heterozygous | Heterozygous | Heterozygous | Heterozygous | Heterozygous | Heterozygous | Heterozygous |
| **Origin** | *De novo* * | *De novo* | *De novo* | *De novo* | *De novo* | *De novo* | *De novo* | *De novo* |
| **Domain** | Coiled-coil | WD40 | WD40 | WD40 | WD40 | WD40 | WD40 | WD40 |
| **References** | Tokita et al., 2018 | Castilla-Vallmanya et al., 2020 | Castilla-Vallmanya et al., 2020; Paprocka et al., 2021 | Paprocka et al., 2021 | Scott et al., 2022 | Castilla-Vallmanya et al., 2020 | This study | Tokita et al., 2018; Castilla-Vallmanya et al., 2020; Accogli et al., 2020; Song et al., 2021 |

\*Ovodonation

## DISCUSSION

Here we provide a detailed description of eleven new TRAF7 syndrome cases, harboring eight different *TRAF7* variants, one of them novel. We also provide a detailed phenotypic description and a comprehensive review of the clinical aspects of this disorder including the previously published cases and propose recommendations for clinical follow-up.

Clinical presentation was mostly consistent with the phenotypic spectrum previously described in TRAF7 syndrome, the leitmotif being the coexistence of four findings: dysmorphic features, developmental delay, growth issues and short stature and cardiac defects.

Despite developmental delay and hypotonia being constant during infancy, some individuals showed normal cognitive function later on. All patients presented good communicative engagement but they all suffered some delay in speech acquisition and language development, with particular impact on expressive language rather than receptive language. As with intellectual disability, language problems were highly variable within the group. While some patients showed a good evolution reaching almost normal communication in adulthood, others remained highly impaired, requiring alternative communications such as signs or pictograms. Considering these results, the use of alternative or augmentative communication methods is recommended for TRAF7 syndrome.

Hearing loss, both conductive and neurosensorial, is also a hallmark that may importantly interfere with communication skills. Therefore it is paramount to perform periodical hearing assessment as indicated in the suggested clinical guidelines (Figure 2) since an early detection may facilitate therapeutic approaches such as tympanic drainages or the use of audiology devices, leading to an improvement in hearing and, consequently, optimizing communication skills.

Autistic traits were present in some of the cases in our cohort, with a particular emphasis on rigidity and lack of cognitive flexibility. However, only two patients fulfilled ASD diagnostic criteria. While in some of the others a formal diagnosis was not reached, the symptomatology is still deserving of attention [26]. Future challenges are to achieve an accurate diagnosis and to set up appropriate intervention and management actions in order to optimize the functional prognosis of the TRAF7 syndrome patients and families. Many of the patients here had a happy and very friendly character although they presented certain difficulties in social skills, with cognitive rigidity, as well as obsession with routines, blockages and tantrums due to frustration. These traits may collectively constitute a distinctive behavioral phenotype in TRAF7 patients and some patients would require follow-up by psychologists or mental health professionals. Previous literature regarding these behavioral traits is scarce but, as it frequently alters the family’s functionality and wellbeing, they should be highlighted [27,28].

While sleep disturbances were not universal or severe among these patients, almost half of the patients present with some kind of affectation, especially referring to the sleep-wake transition. Of note, increased sleepiness, observed here in three patients, was also mentioned in two previously published subjects [1]. Sleep assessment and follow-up is relevant and should be taken into consideration in TRAF7 patients as sleep disturbances lead to a decline in cognitive performance and behavior.

Regarding endocrine characteristics, although short stature is an early sign, at birth it may not be striking enough unless accompanied by other signs. During childhood, short stature becomes more notorious and is usually accompanied by proportionate or low weight. Due to the scarcity of data, it is unclear whether this is genetically determined stature or it may be explained by a hormonal deficiency during puberty.

Hormonal factors have not been adequately studied in TRAF7 cases, and overall somatometric data is insufficient, but in spite of the small number of individuals taken into consideration, our observations suggest that this deficiency worsened after puberal age (Figure 1). It is important to take this factor into account in future reviews of patients with TRAF7 syndrome since it may have therapeutic implications, as it has been shown in some genetic syndromes combining endocrine and neurological disturbances [29].

As in the previously described patients, structural congenital heart defects were present in a majority of TRAF7 patients. These defects trigger the most severe complications, especially during their early life. In our cohort, patent ductus arteriosus was the most prevalent manifestation together with a combination of severe cardiac, valvular and vessel pathologies including life-threatening abnormalities such as hypoplastic aortic arch leading to cardiac insufficiency in four patients in the cases presented here. The involvement of TRAF7 in endothelial tissues, and particularly regarding vascular complications, has been recently demonstrated in a murine model [14].

As in previous reports, patients here described with the particular gestalt of the TRAF7 syndrome patients [2] especially on the upper face, including blepharophimosis, ptosis, hypertelorism and a wide or flat nasal bridge. Additional facial features are a bulbous nasal tip and dysplastic or low set ears. Further revision of previously published pictures together with the cases presented here highlights that arched eyebrows are also relatively common in this syndrome.

A novel variant has been described in this work, p.(Arg653Leu), identified in Patient 7. It is a missense variant affecting the Arg653 residue, where other missense somatic changes associated with meningioma tumors have been described. In fact, p.Arg653Gln and p.Arg653Pro are recurrent variants in those tumors.

This is not the first time that germline changes associated with TRAF7 syndrome and somatic changes in meningioma have been identified in the same residue, as is the case with the germline variant p.(Leu519Phe) associated with TRAF7 disorder and somatic variants p.(Leu519Pro) and p.(Leu519Arg), described in meningioma tumors. This apparent division where some missense changes in germline lead to TRAF7 syndrome whereas similar changes in somatic tissue give rise to tumor conditions is something that needs to be clarified. Further, Kaidonis et al. (2022) recently described an individual with bilateral optic nerve sheath meningiomas and TRAF7 syndrome who had the c.1921C>T p.(Arg641Cys) variant in mosaicism, previously associated with meningiomas [6] . Two additional individuals with TRAF7 syndrome have been reported to develop meningiomas [1,2]. This raises the question of whether individuals with germline variants in TRAF7 may have an increased risk of developing malignancies such as meningiomas. We believe it is important to keep this fact in mind as new evidence on this relationship is gathered.

Diagnosis of TRAF7 syndrome might be hampered by the presence of features overlapping with other syndromes, which resulted in different initial diagnosis orientations. FAT1, *KAT6B*, RASopathies, or connective tissue disorders were considered as likely players in initial diagnosis. KAT6B-related disorder Ohdo syndrome (SBBYSS; MIM #603736) has common characteristics with TRAF7 syndrome, such as motor and speech delay, digital anomalies, cardiac defects, and craniofacial features, including blepharophimosis and ptosis [30]. Also, connective disorders and RASopathies (e.g., Costello syndrome) are typically suspected in TRAF7 patients migth be due to a correlation in the pathomechanisms of both RASopathies and TRAF7 syndrome involving the regulation of RAS/MAPK signaling pathways [1,5].

This new cohort of TRAF7 syndrome patients expands the phenotypic spectrum of this disorder that presents with variable but recognizable features, particularly referring to behavior or sleep disturbances. In this work, we address growing issues that need more attention in future studies such as autistic traits, short stature, and hearing loss. An in depth revision led to the first proposal of a based-on-evidence schedule to help in the clinical management and monitoring of TRAF7 syndrome patients.

## Data Availability

All data produced in the present study are available upon reasonable request to the authors

## ACKNOWLEDGEMENTS

We would like to thank all the patient’s and their families for their collaboration.

## FUNDING

APP is recipient of an FPU fellowship from Spanish Ministerio de Universidades.

ESG is supported by CIBERER through the project IMPaCT-GENóMICA (IMP/00009) ISCIII from Ministerio de Sanidad, Madrid, Spain cofunded by European Union.

MCP is supported by a Carmen de Torres fellowship from IRSJD.

Funding was provided by grants PID2019-107188RB-C21 and PID2022-141461OB-I00 from Spanish MCIN/AEI/ 10.13039/501100011033; and grant 2021SGR-01093 from AGAUR-Generalitat de Catalunya.

Funding sources were not involved in the study design, collection, analysis, and interpretation of data, writing of the report or the publication of the article.

## Competing Interests

The authors declare no competing interests

## Notes

### Competing Interest Statement

The authors have declared no competing interest.

### Funding Statement

This study was funded by 
- FPU fellowship from Spanish Ministerio de Universidades.
- CIBERER through the project IMPaCT-GENoMICA (IMP/00009) ISCIII from Ministerio de Sanidad, Madrid, Spain cofunded by European Union.
- Carmen de Torres fellowship from IRSJD.
Funding was provided by grants PID2019-107188RB-C21 and PID2022-141461OB-I00 from Spanish MCIN/AEI/ 10.13039/501100011033; and grant 2021SGR-01093 from AGAUR-Generalitat de Catalunya.
Funding sources were not involved in the study design, collection, analysis, and interpretation of data, writing of the report or the publication of the article.

### Author Declarations

Comision de Bioetica de la Universidad de Barcelona gave ethical approval for this work.

