## Supplemental Figure 1 for "Expanding the phenotypic spectrum of TRAF7 syndrome: report of eleven new cases and literature review"

### Slide 1
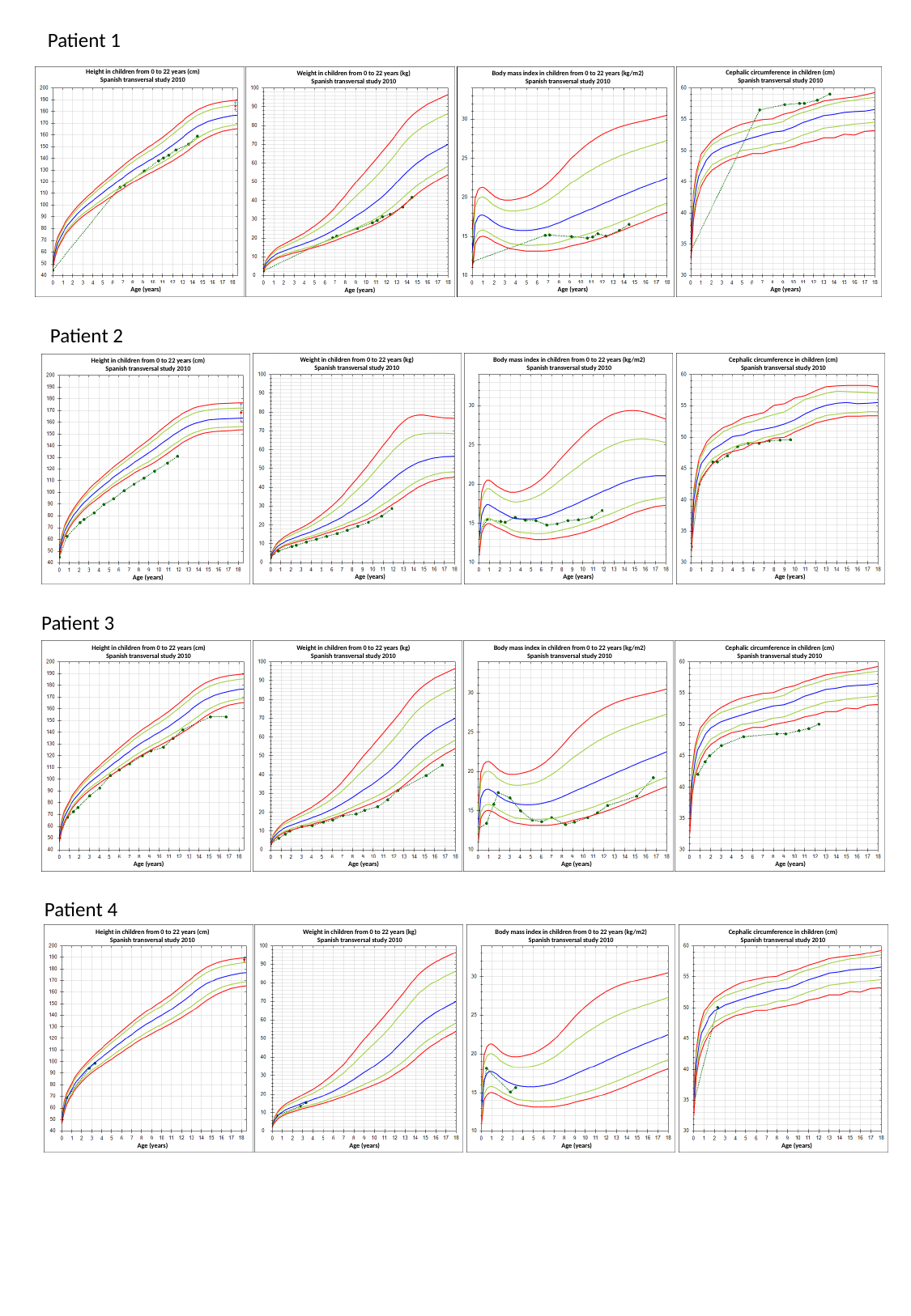

Patient 1
Height in children from 0 to 22 years (cm)
Spanish transversal study 2010
Age (years)
Age (years)
Age (years)
Age (years)
Weight in children from 0 to 22 years (kg)
Spanish transversal study 2010
Body mass index in children from 0 to 22 years (kg/m2)
Spanish transversal study 2010
Cephalic circumference in children (cm)
Spanish transversal study 2010
Patient 2
Height in children from 0 to 22 years (cm)
Spanish transversal study 2010
Age (years)
Age (years)
Age (years)
Age (years)
Weight in children from 0 to 22 years (kg)
Spanish transversal study 2010
Body mass index in children from 0 to 22 years (kg/m2)
Spanish transversal study 2010
Cephalic circumference in children (cm)
Spanish transversal study 2010
Patient 3
Height in children from 0 to 22 years (cm)
Spanish transversal study 2010
Age (years)
Age (years)
Age (years)
Age (years)
Weight in children from 0 to 22 years (kg)
Spanish transversal study 2010
Body mass index in children from 0 to 22 years (kg/m2)
Spanish transversal study 2010
Cephalic circumference in children (cm)
Spanish transversal study 2010
Patient 4
Height in children from 0 to 22 years (cm)
Spanish transversal study 2010
Age (years)
Age (years)
Age (years)
Age (years)
Weight in children from 0 to 22 years (kg)
Spanish transversal study 2010
Body mass index in children from 0 to 22 years (kg/m2)
Spanish transversal study 2010
Cephalic circumference in children (cm)
Spanish transversal study 2010

### Slide 2
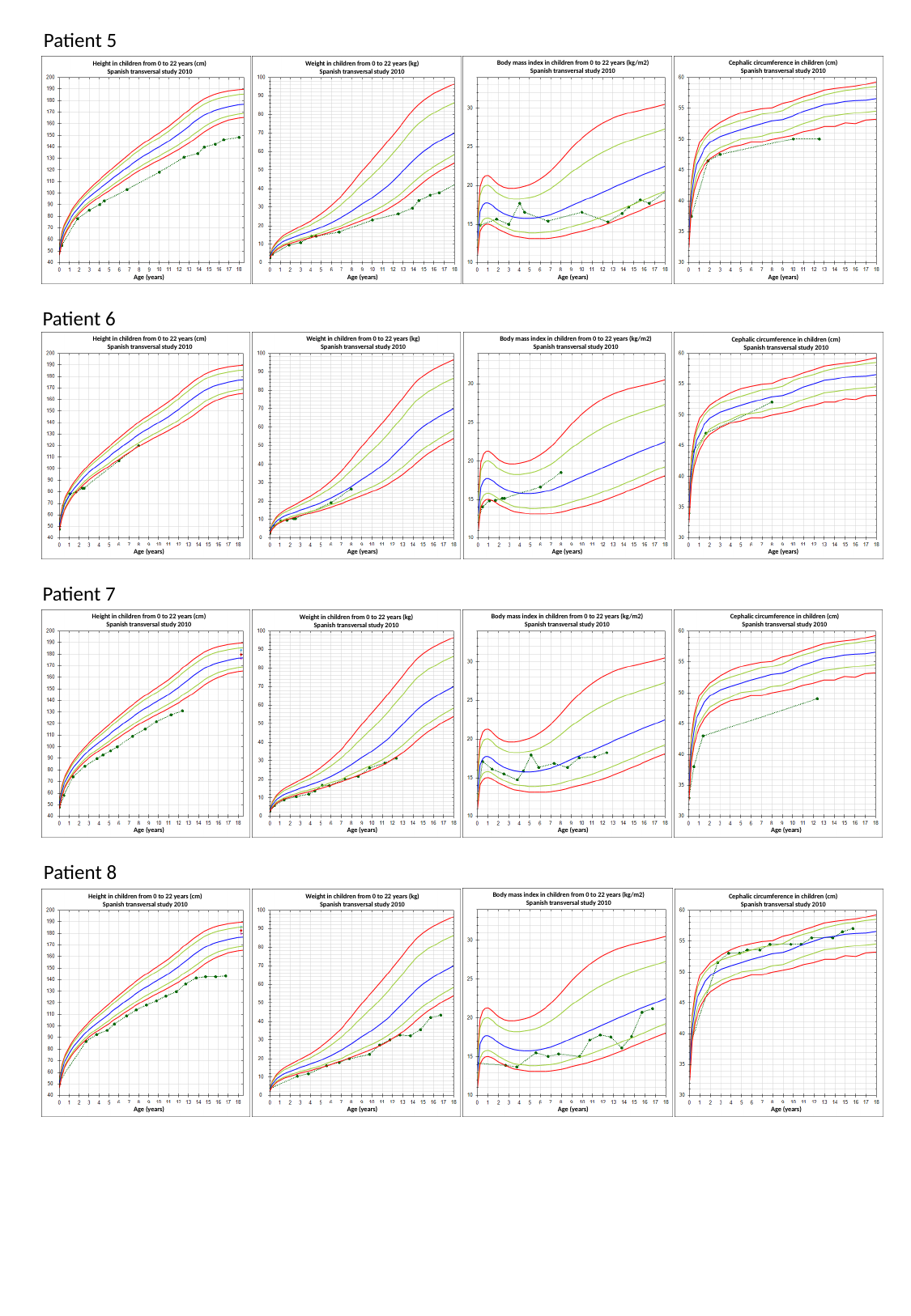

Patient 5
Height in children from 0 to 22 years (cm)
Spanish transversal study 2010
Age (years)
Age (years)
Age (years)
Age (years)
Weight in children from 0 to 22 years (kg)
Spanish transversal study 2010
Body mass index in children from 0 to 22 years (kg/m2)
Spanish transversal study 2010
Cephalic circumference in children (cm)
Spanish transversal study 2010
Patient 6
Height in children from 0 to 22 years (cm)
Spanish transversal study 2010
Age (years)
Age (years)
Age (years)
Age (years)
Weight in children from 0 to 22 years (kg)
Spanish transversal study 2010
Cephalic circumference in children (cm)
Spanish transversal study 2010
Body mass index in children from 0 to 22 years (kg/m2)
Spanish transversal study 2010
Body mass index in children from 0 to 22 years (kg/m2)
Spanish transversal study 2010
Patient 7
Height in children from 0 to 22 years (cm)
Spanish transversal study 2010
Age (years)
Age (years)
Age (years)
Age (years)
Height in children from 0 to 22 years (cm)
Spanish transversal study 2010
Weight in children from 0 to 22 years (kg)
Spanish transversal study 2010
Body mass index in children from 0 to 22 years (kg/m2)
Spanish transversal study 2010
Cephalic circumference in children (cm)
Spanish transversal study 2010
Patient 8
Height in children from 0 to 22 years (cm)
Spanish transversal study 2010
Age (years)
Age (years)
Age (years)
Age (years)
Weight in children from 0 to 22 years (kg)
Spanish transversal study 2010
Cephalic circumference in children (cm)
Spanish transversal study 2010
Body mass index in children from 0 to 22 years (kg/m2)
Spanish transversal study 2010
Body mass index in children from 0 to 22 years (kg/m2)
Spanish transversal study 2010

### Slide 3
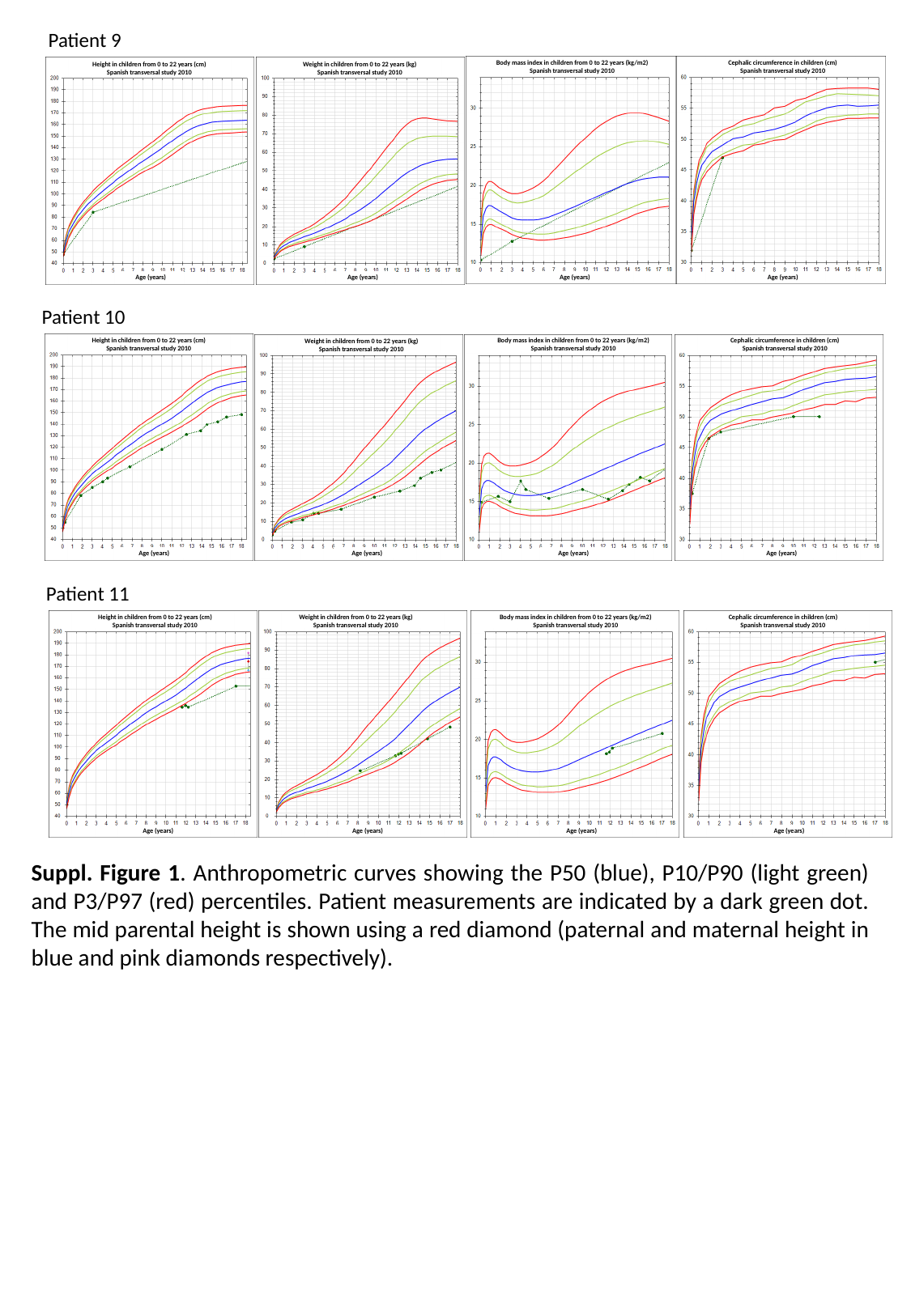

Patient 9
Height in children from 0 to 22 years (cm)
Spanish transversal study 2010
Age (years)
Age (years)
Age (years)
Age (years)
Weight in children from 0 to 22 years (kg)
Spanish transversal study 2010
Body mass index in children from 0 to 22 years (kg/m2)
Spanish transversal study 2010
Cephalic circumference in children (cm)
Spanish transversal study 2010
Patient 10
Height in children from 0 to 22 years (cm)
Spanish transversal study 2010
Age (years)
Age (years)
Age (years)
Age (years)
Weight in children from 0 to 22 years (kg)
Spanish transversal study 2010
Body mass index in children from 0 to 22 years (kg/m2)
Spanish transversal study 2010
Cephalic circumference in children (cm)
Spanish transversal study 2010
Patient 11
Height in children from 0 to 22 years (cm)
Spanish transversal study 2010
Age (years)
Age (years)
Age (years)
Age (years)
Weight in children from 0 to 22 years (kg)
Spanish transversal study 2010
Body mass index in children from 0 to 22 years (kg/m2)
Spanish transversal study 2010
Cephalic circumference in children (cm)
Spanish transversal study 2010
Suppl. Figure 1. Anthropometric curves showing the P50 (blue), P10/P90 (light green) and P3/P97 (red) percentiles. Patient measurements are indicated by a dark green dot. The mid parental height is shown using a red diamond (paternal and maternal height in blue and pink diamonds respectively).
